# HIV-infected patients on combined antiretroviral treatment had similar level of arterial stiffness to the patients with ST-segment elevation myocardial infarction

**DOI:** 10.1101/2021.01.08.21249466

**Authors:** Tomasz Mikuła, Magdalena M. Suchacz, Michał Peller, Paweł Balsam, Łukasz Kołtowski, Renata Główczyńska, Krzysztof J. Filipiak, Grzegorz Opolski, Alicja Wiercińska-Drapało

**Author notes:** Corresponding author: Tomasz Mikuła, Department of Infectious and Tropical Diseases and Hepatology, Medical University of Warsaw, 01-201 Warsaw, Wolska 37, Poland.

## Abstract

**Purpose:** The cardiovascular disease has become very common among HIV-infected patients. The aim was to compare the arterial stiffness and the endothelial dysfunction in HIV-infected patients to non-HIV-infected patients in week 4 after ST-segment elevation myocardial infarction (STEMI).

**Methods:** The arterial stiffness was calculated by Endo-PAT 2000(ITAMAR®) and the endothelial function by Peripheral Arterial Tonometry(PAT®). The correct endothelial function was defined for natural logarithm of reactive hyperaemia index (lnRHI)>0.51. Arterial stiffness was assessed as the AI and corrected for heart rate of 75 bpm (AI@75).

**Results:** Sixty-three patients were recruited to this study, n=34 patients with HIV infection (n=18 on cART) and n=29 HIV-negative patients after recent STEMI. No statistically significant differences for AI and AI@75 were found in STEMI and in HIV on cART group. We observed p<0.05 for AI and AI@75 for patients without cART compared to STEMI and on cART patients. The observed lnRHI results were significantly different p<0.05 in STEMI and on cART patients. We get similar endothelial dysfunction p>0.05 for patients without cART compared to STEMI and on cART patients.

**Conclusions:** Assessing cardiovascular risk also with non-invasive methods among HIV-infected patients is very important especially in HIV-patients on cART. Endothelial dysfunction is connected with HIV infection and can be similar for STEMI and HIV-infected without cART.

## Introduction

Effective combined antiretroviral therapy (cART) has significantly improved life expectancy in HIV-infected persons. Consequently, the age-related co-morbidities, as cardiovascular disease, have become more common in this population. HIV is an independent risk factor for cardiovascular disease, and people with HIV infection have a higher risk of myocardial infarction than those who are HIV-negative [1,2]. There are no known mechanisms related to the influence of HIV-1 proteins on the vascular endothelium, however, some synergy has been demonstrated between selected proinflammatory cytokines and some viral proteins. For instance, TNF-alpha and IL-6 are secreted in response to the mononuclear cell activation or to the direct action of HIV proteins such as gp 120, nef and tat [3,4]. Moreover, Nef protein, inhibits the activity of adenosine triphosphate binding cassette A1 transporter (ABCA1), what significantly reduces the outflow of cholesterol from macrophages and increases the risk of atherosclerosis by negative correlation with the thickness of the intima-media in the carotid arteries regardless of HDL (High-density lipoprotein) cholesterol level [5,6]. Nadel et al. compared HIV-infected to non-HIV-infected patients over a median period of 38 months showing an increased risk of acute coronary syndrome without ST-segment elevation (NSTEMI) in HIV-infected patients. Moreover, they showed that HIV-infected patients had more severe coronary atherosclerosis in CT angiography and higher rates of NSTEMI compared to uninfected patients [7]. The attention was also focused on the significant effect of cART on the lipid metabolism and the increased cardiovascular risk during the use of certain drug groups, especially protease inhibitors (PIs), non-nucleoside reverse transcriptase inhibitors - NNRTIs (efavirenz), and nucleoside reverse transcriptase inhibitors - NRTIs (zidovudine) [8,9].

Evaluating cardiovascular risk among HIV-infected people is not an easy assignment. The Framingham scale is being used far less frequently to calculate the 10-year risk of myocardial infarction, as the cardiovascular risk in this group of patients is often underestimated. Moreover, the importance of cART treatment, interaction between drugs and undesirable drug effects are underlined [10,11]. There are many invasive and non-invasive methods to assess endothelial function and arterial stiffness. One of the non-invasive techniques is Reactive Hyperaemia Peripheral Arterial Tonometry (RH-PAT) which allows to evaluate the peripheral microcirculation vessels. The correct endothelial function was defined for natural logarithm of Reactive Hyperaemia Index (lnRHI)>0.51 [12]. Using the system with a pair of plethysmographic sensors of Peripheral Arterial Tonometry (PAT®) it is possible to evaluate changes in the peripheral vascular arteria tone - Augmentation Index (AI) - and normalised it for heart rate of 75 bpm (beats per minute) - AI@75 [13,14]. AI is a non-invasive method of estimating arterial stiffness.

The aim of this study was to compare the arterial stiffness and the endothelial dysfunction in HIV-infected patients (with or without cART) to non-HIV-infected patients in week 4 after ST-segment elevation myocardial infarction (STEMI).

## Materials and methods

We evaluated and compared the arterial stiffness and the endothelial dysfunction in adult HIV-infected patients with non-HIV-infected persons after STEMI. The only exclusion criterion for both groups was having experienced myocardial infarction in anamnesis.

On the visit day, each HIV-infected patient underwent a basic medical examination. The patients were questioned on the minimum amount of smoked cigarettes as well as other accompanying diseases. People who smoked a minimum of 20 cigarettes a day for a minimum of 5 years were considered. The Body Mass Index (BMI) was assessed and data from basic biochemical tests were collected. Next, the arterial stiffness was calculated by using the Endo-PAT 2000 (ITAMAR®) and the endothelial function by using Peripheral Arterial Tonometry (PAT®). The correct endothelial function was defined for natural logarithm of reactive hyperaemia index (lnRHI)>0.51. Arterial stiffness was assessed as the Augmentation Index (AI) and AI values were corrected for heart rate using an arbitrarily defined reference heart rate of 75 bpm (AI@75).

Among HIV-negative patients after STEMI, similar procedures were carried out in the 4th week of admission to the hospital with the exception of laboratory tests that were performed at the time of acute coronary syndrome (ACS).

All participants signed a written informed consent. The study was approved by the Bioethics Committee.

All categorical variables were presented as percentages. To assess distributions of continuous variables Shapiro-Wilk test was used. For all analysed continuous variables normal distributions were found. Continuous variables were presented as mean values and standard deviations. Differences between subgroups were calculated with Fisher’s exact test and Student’s t-test, respectively for categorical and continuous variables. To reduce the age as a confounding variable, propensity score matching was used, with the 1:1 ratio for numbers of patients in each subgroup. Tests were considered significant for p-values <0.05. Statistical analysis was performed using SAS® software, version 9.4.

## Results

Sixty-three patients were recruited to this study, n=34 patients with HIV-infection (n=18 of them on cART treatment) and n=29 HIV-negative patients after experiencing STEMI. All results obtained from both groups are presented in Table 1.

**Table 1.**
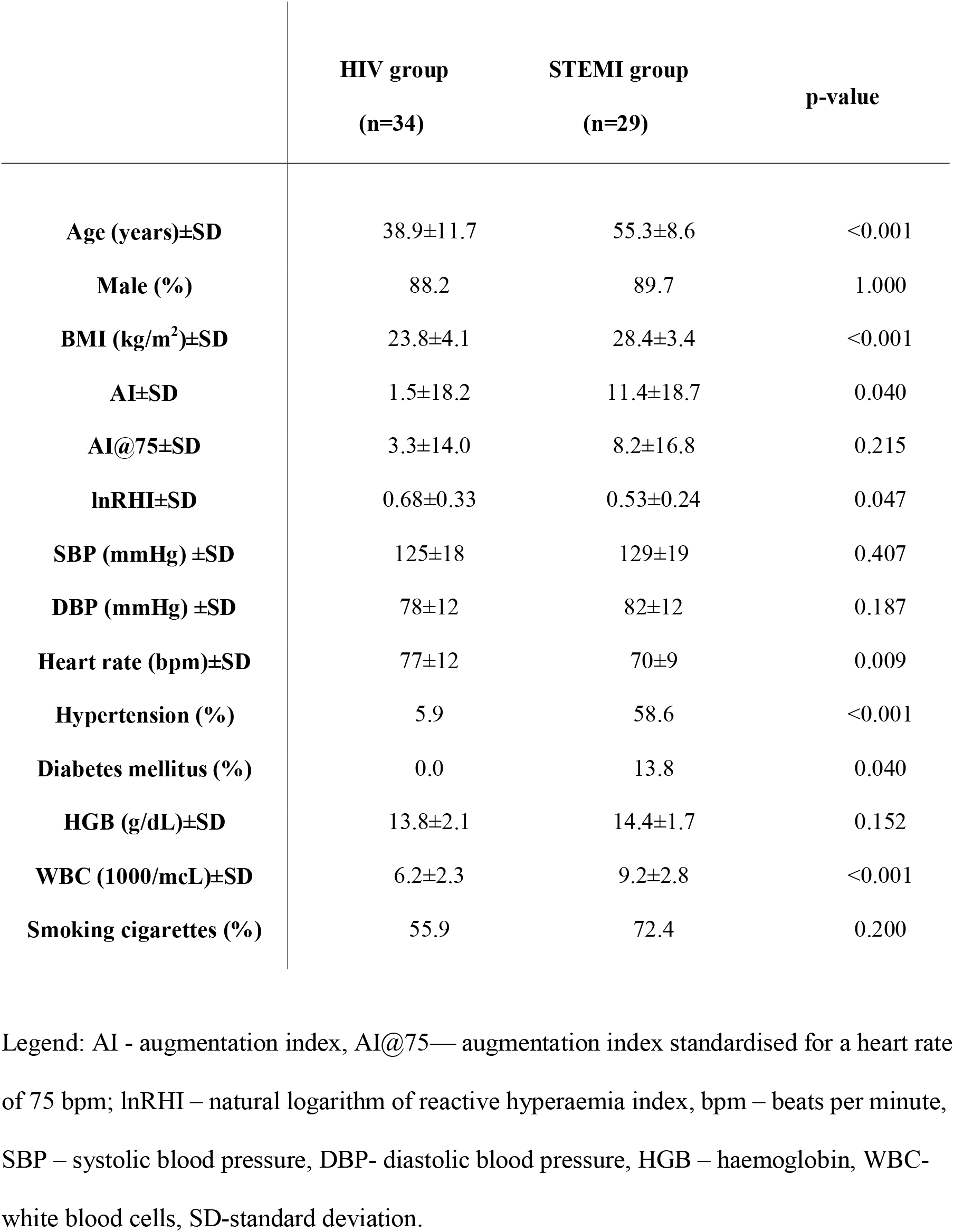
Baseline patients’ characteristics.

We compared AI, AI@75, and lnRHI, for three study groups: STEMI, HIV group on cART and HIV group without cART. All results were presented in Table 2.

**Table 2.** Comparison of AI, AI@75, lnRHI in STEMI and HIV group on and without cART (naïve).

|  | STEMI<br>group<br>(n=29) | HIV group<br>on cART<br>(n=18) | HIV group<br>without<br>cART<br>(n=16) | p-value |
| --- | --- | --- | --- | --- |
| **AI**<br>**(min-max)** | 11<br>(3-19) | 7<br>(-4-14) | -7<br>(-19-1) | STEMI vs on cART: 0.584<br>STEMI vs without cART <**0.001**<br>On cART vs without cART: **0.002** |
| **AI@75**<br>**(min-max)** | 8<br>(1-14) | 8<br>(-1-13) | -3<br>(-10-4) | STEMI vs on cART: 0.861<br>STEMI vs without cART: **0.013**<br>On cART vs without cART: **0.025** |
| **lnRHI**<br>**(min-max)** | 0.47<br>(0.37-0.67) | 0.75<br>(0.45-0.98) | 0.62<br>(0.38-0.85) | STEMI vs on cART: **0.032**<br>STEMI vs without cART: 0.286<br>On cART vs without cART: 0.301 |

Legend: AI - augmentation index, AI@75— augmentation index standardised for a heart rate of 75 bpm; lnRHI – natural logarithm of reactive hyperaemia index.

Subsequently, from the received data, we created two groups matched in terms of age and parity (1:1) to obtain the data presented in Table 3.

**Table 3.** The groups matched in terms of age and parity (1:1).

|  | **HIV group**<br>**(n=20)** | **STEMI group**<br>**(n=20)** | **p-value** |
| --- | --- | --- | --- |
| **Age (years)±SD** | 45.8±10.3 | 50.8±5.8 | 0.069 |
| **Male (%)** | 90.0 | 85.0 | 1.000 |
| **AI±SD** | 6.9±19.3 | 12.0±21.1 | 0.435 |
| **AI@75 bpm±SD** | 8.3±13.6 | 9.1±18.8 | 0.879 |
| **lnRHI±SD** | 0.71±0.34 | 0.52±0.24 | 0.057 |

Legend: AI - augmentation index, AI@75— augmentation index standardised for a heart rate of 75 bpm; lnRHI – natural logarithm of reactive hyperaemia index, bpm – beats per minute, SD-standard deviation.

The cART treatment in HIV group contained four classes of drugs. The average duration of therapy was approximately 43 months (3.6 years). The numbers of patients with particular cART drug are shown in Table 4.

**Table 4.** The numbers of patients with particular cART drug class.

| **Antiretroviral therapy** | **(n)** |
| --- | --- |
| Nucleoside Reverse Transcriptase Inhibitors (NRTIs) | 13 |
| Protease Inhibitors (PIs) | 14<sup>a</sup> |
| Non-nucleoside Reverse Transcriptase Inhibitors (NNRTIs) | 4<sup>b</sup> |
| Integrase Strand Transfer Inhibitors (INSTIs) | 4<sup>c</sup> |

<sup>a</sup> All patients with ritonavir as a booster
<sup>b</sup> 3 patients with efavirenz and 1 with nevirapine
<sup>c</sup> All patients with raltegravir

Among 18 patients on cART, 5 (27.7%) had endothelial dysfunction, and 10 (55.5%) smoked cigarettes. In the group of 16 patients without cART, 7 (43.7%) had endothelial dysfunction and 9 (62.5%) smoked cigarettes.

## Discussion

The arterial stiffness is a predictor of endothelial dysfunction in subclinical patients. The determination of the augmentation index (AI) is a non-invasive method of its estimation. AI has been already used as a marker of subclinical atherosclerotic disease in people living with HIV [15]. We have been previously showed that HIV infection and cART had negative influence on arterial stiffness [16].

In current study, we compared AI results from HIV-infected patients with or without cART with persons after myocardial infarction. As a result, we showed that AI was significantly higher in STEMI group than in HIV-infeted group especially without cART. No statistically significant difference of arterial stiffness were found between STEMI and HIV-infected patients on cART before and after AI@75 standardisation. In our study we also showed that HIV-infected without cART had significantly lower AI and AI@75 results in comparison to cART-treated and STEMI group.

Moreover, after matching 1:1 the examined groups in accordance to age and number, we confirmed that the arterial stiffness in HIV-infected group is similar to STEMI patients, what may be an indirect proof of the high advancement of cardiovascular changes in this group of patients.

Our HIV-infected patients were significantly younger and had lower BMI than patients with STEMI. Futhermore, in our STEMI group a high percentage of patients with diabetes and hypertension (13.8% and 58.6%, respectively) was observed in comparison to HIV group (0% and 5.9%, respectively). These differences were statistically significant and showed that there are the other factors, apart from the well-known environemental and genetical factors, which probably enhance arterial stiffness in HIV-positive patients. Effective cART has significantly improved life expectancy in people living with HIV-infection. On the other hand, cART influence on the increase of cardiovascular risk has been confirmed. CART drugs, especially ritonavir boosted protease inhibitors (PIs/r), significantly impact on serum lipids and enhance atherosclerotic lesions [17,18]. There are several studies concerning HIV and cART influence on the arterial stiffness augmentation. Their results are disagreeing as some studies confirmed this association and some others did not [19]. In effect, our results may confirm the negative cART (especially containing PIs) influence on the arterial stiffness and on the subclinical atherosclerotic disease development in HIV-infected persons.

Besides, we analysed the endothelial function by using Peripheral Arterial Tonometry and Reactive Hyperaemia Index (RHI). In our study, observed lnRHI results were significantly lower in STEMI group in comparison to HIV-infected patients what may suggest more intense endothelial dysfunction in the group of persons after myocardial infarction. This trend was also observed after analysis of groups matched in terms of age and number. The biggest significant difference in the degree of endothelial dysfunction was seen between STEMI group and cART-treated HIV-infected patients what may suggest the protective cART effect on endothelial function. However, we did not show the differences between lnRHI results in HIV-infected patients with and without cART.

The importance of WBC (White Blood Cells) count in patients with myocardial infarction has been described in many studies. In patients with STEMI and NSTEMI, the WBC number is important in the first hours of an acute cardiovascular episode as a prognostic marker for patients undergoing primary percutaneous coronary intervention [20,21]. In our study, we observed a statistically significant higher number of WBC in the STEMI group in comparison to HIV-infected patients. On the one hand, it could confirm the observation that WBC is augmented in persons after myocardial infarction. On the other hand, it may be the effect of leukopenia often observed among HIV-infected patients.

Cigarette smoking is one of the main factors that increase endothelial changes and is also an independent cardiovascular risk factor [22,23]. In our study we observed a high percentage of patients who smoked cigarettes in both groups. We did not show a statistically significant difference in rate of smoking persons in studied groups. For this reason, the risk of coronary vascular injury caused by smoking was similar in all studied patients. Futhermore, HIV infection is an independent risk factor for cardiovascular disease development, hence the combination of these two factors is particularly dangerous for HIV-infected patients.

One limitation of this study is a relatively small number of participants however we were able to show that cART use may have subsequently negative influence on arterial stiffness as recent myocardial infarction. Nevertheless, our findings should be confirmed in larger studies. In addition, among HIV-infected patients, there was no one who started hypolipidemic treatment before being included to the study. In the STEMI group no one previously used hypolipidemic therapy. However, when STEMI occurred, everyone received this treatment. According to the study methods, the assessment of arterial stiffness and endothelial dysfunction in the STEMI group was performed in week 4 after acute coronary syndrome. However, it seems that after such a short period of time, the hypolipidemic treatment should have an unimportant effect on analysed parameters.

## Conclusions

In summary, we report that HIV-infected on cART may have similarly high values of arterial stiffness as non-HIV-infected persons in week 4 after STEMI. Assessing cardiovascular risk is very important especially in infected patients on cART therapy. Modification of risk factors should be basic element of care for patients infected with HIV. More studies are required to assess our observation concerning arterial stiffness and elevated risk of cardiovascular disease development in this group of patients, independently of age, smoking status and concomitant chronic diseases.

Among our knowledge, this study is the first comparing the arterial stiffness and the endothelial dysfunction between STEMI patients not infected with HIV to HIV-infected patients with and without cART.

## Data Availability

We confirm the statement regarding the availability of all data referred to in the manuscript.

